# The impact of social and structural support on anxiety among academics in health professions fields

**DOI:** 10.1101/2025.10.13.25337911

**Authors:** Anietie Andy, Natasha Tonge, Praise-EL Michaels, Marina K. Holz

## Abstract

Academic faculty in the health professions are especially susceptible to anxiety-inducing factors as a consequence of their demanding professional lives. The COVID-19 pandemic placed special attention on the experiences of anxiety and stress among health professionals and academics; however, there is a lack of studies of the impact of specific types of social modifiers of anxiety in health professions faculty. To address this critical gap, we administered the Generalized Anxiety Disorder (GAD-7) questionnaire to 549 self-identified academic faculty in health professions. Of those surveyed, 8.1% reported severe levels of anxiety and 19.6% reported moderate levels, totaling 27.7% with moderate-to-severe symptoms. Our results revealed that academic rank significantly predicted anxiety levels, with assistant professors (both tenure-track and non-tenure track) experiencing significantly higher anxiety than associate and full professors or adjunct or part-time faculty. Age demonstrated strong protective effects, while women and gender minorities reported elevated anxiety levels compared to men. Having close family relationships and academic parents were both associated with significantly lower anxiety, while spouse occupation in non-academic sectors provided protective effects. Social responsibilities of having children or larger family size did not significantly impact anxiety levels. Our results showed that post-pandemic anxiety levels remain notable and potentially impairing among academic health professionals, and that structural and social supports, and demographic factors can act as buffers against anxiety. Our study can inform future efforts by institutions to support academic health professional faculty mental health.

## Introduction

Anxiety, characterized by fear or worry about the future^1^ is highly comorbid with other mental health conditions such as depression at rates of over 75%^2^. Anxiety and vulnerability to anxiety may increase risk for other health conditions^3^. Globally, anxiety presents an unaddressed mental health need and a significant public health burden^4^. Several studies have investigated mental health and anxiety expression in health professions trainees, such as medical and doctoral students^5-7^. Our prior work examined prevalence and modifiers of anxiety among academic faculty, highlighting the disproportionate anxiety burden on early-career academics and establishing that familial and parental relationships can ease anxiety in this population^8^.

However, there exists a gap in knowledge regarding the needs and mental health burdens of health professions faculty. Health professions faculty lead complex professional lives and are uniquely susceptible to anxiety-inducing factors. Faculty members integrate multiple demanding responsibilities across clinical practice, research, teaching, and administrative duties, leading to chronic role overload and significant time management challenges^9^. Career progression requirements on competitive promotion and tenure track add a layer of instability and uncertainty^10^. Academic success hinges on securing research funding, publish-or-perish academic culture, producing high-impact research and navigating rigorous peer review processes, further exacerbating anxiety. Financial pressures add another layer of stress, including relatively low compensation compared to clinical practice, and ongoing student loan burdens. The hierarchical institutional structures, combined with cultural expectations of persistent high performance, create an environment where mental health is susceptible to being marginalized. In contrast, adjunct and part-time faculty, while facing their own challenges including limited benefits and job security, have reduced expectations for research productivity and advancement. Their roles are often more clearly defined and may involve fewer competing demands than full-time academic positions.

Emerging research suggests that anxiety expression in academic settings varies significantly across demographic groups^8^. Women and gender minorities face unique stressors in traditional academic environments that may not be structured to accommodate diverse gender identities. Younger academics enter a more competitive and uncertain academic job market than previous generations, with increased pressure for productivity and decreased availability of tenure-track positions. The financial burden of extended education, including substantial student loan debt, may compound anxiety among younger faculty members. Racial and ethnic minorities in academic medicine face additional systemic barriers that can contribute to anxiety, including underrepresentation, bias in hiring and promotion, and limited access to mentorship from individuals with similar backgrounds. The intersection of multiple minority identities may create compounded stressors. Therefore, there exists a critical need to study the expression of anxiety among health professions faculty, to inform tailored interventions that can take into account both individual and institutional factors, and to create durable and meaningful institutional support systems.

## Methods

### Survey Design and Participants

We conducted a cross-sectional survey of faculty from health professions programs from 62 higher education and medical institutions. Participants were recruited through professional associations, institutional directories, and snowball sampling, and contacted via email with an invitation to complete survey instruments via Qualtrics. A total of 549 self-identified academic health professionals completed the survey. The survey was approved by the Howard University Institutional Review Board. Participants were not compensated for their participation.

### Measures

#### Demographic Variables

Participants provided comprehensive demographic information including age range (18-24, 25-34, 35-44, 45-54, 55-64, 65-74, 75 or older), race/ethnicity (White, Asian or Asian American, Black or African American, Hispanic/Latino, Biracial/multiracial/mixed race, American Indian or Native American), and gender identity (female, male, or non-binary/third gender).

#### Professional Characteristics

Participants with an academic appointment in the health professions, defined as schools/programs of medicine, dentistry, pharmacy, physical therapy, indicated their institution type (research-intensive academic or medical center, public 4-year university or college, private 4-year university or college, Historically Black College or University (HBCU) or Hispanic-serving institution, or liberal arts college), annual income, and detailed academic rank classification (Assistant Professor tenure or non-tenure track, Associate Professor tenure or non-tenure track, Full Professor tenure or non-tenure track, Adjunct or part-time faculty).

#### Family and Social Support Variables

Family structure was assessed through marital status, number of children, and household size. Social support measures included the presence of close family relationships (yes/no), whether family lived nearby (yes/no), and academic family background assessed by whether participants had at least one parent who was or is an academic faculty member (yes/no). Spouse/partner occupation was categorized as academia, industry/business/for-profit sectors, K-12 education, medical or healthcare, military, retired, or unemployed/unable to work.

#### Anxiety Assessment

Generalized anxiety symptoms were measured using the GAD-7 (Generalized Anxiety Disorder 7-item scale), a well-validated instrument for assessing anxiety severity in clinical and research settings^11^. The GAD-7 produces scores ranging from 0-21, with established cut-points for mild (5-9), moderate (10-14), and severe (15-21) anxiety levels. Scores of 10 or above indicate potential functional impairment from anxiety symptoms.

#### Data Analytic Plan

Descriptive statistics were calculated for all variables using R/RStudio, and regression analyses were conducted using Stata SE version 17 statistical software. Multiple regression analysis was conducted to examine predictors of GAD-7 total scores, with categorical variables entered as factor variables using appropriate reference groups. The dependent variable was “gad_total”, a count variable representing the total GAD-7 score for each respondent. Given the count nature of the dependent variable and evidence of overdispersion (variance exceeding the mean), a negative binomial regression model was employed. Two models were estimated. Model 1 included only academic rank as a predictor, and Model 2 included academic rank along with demographic covariates. Model assumptions were tested, and model fit was assessed using the likelihood ratio chi-square test, pseudo R^2^, and log-likelihood values. The dispersion parameter alpha was estimated to confirm the appropriateness of the negative binomial specification. Coefficients were exponentiated to derive Incidence Rate Ratios (IRRs), which represent the multiplicative change in the expected count of gad_total associated with each predictor. The final model included independent variables academic rank (categorical), presence of a close family relationship (binary), having an academic parent (binary) and age group (categorical), with statistical significance set at *p* < 0.05.

## Results

Overall, 27.7% of respondents had moderate to severe anxiety levels, with GAD-7 score significantly influenced by career stage, age, gender identity, and family factors.

### Academic and Professional Factors

Academic rank emerged as a significant predictor of anxiety severity (Figure 1). Of all respondents, 41.7% were assistant professors, which was identified as the highest-anxiety group. Compared to adjunct or part-time faculty (reference group), assistant Professors in non-tenure track positions reported higher GAD-7 scores, as did tenure-track assistant professors. Associate professors in both non-tenure track and tenure-track positions showed elevations in GAD-7 scores, while full professors showed no significant differences from the reference group. This suggests that junior and mid-career faculty may experience elevated anxiety levels relative to their senior or part-time counterparts.

**Figure 1.**
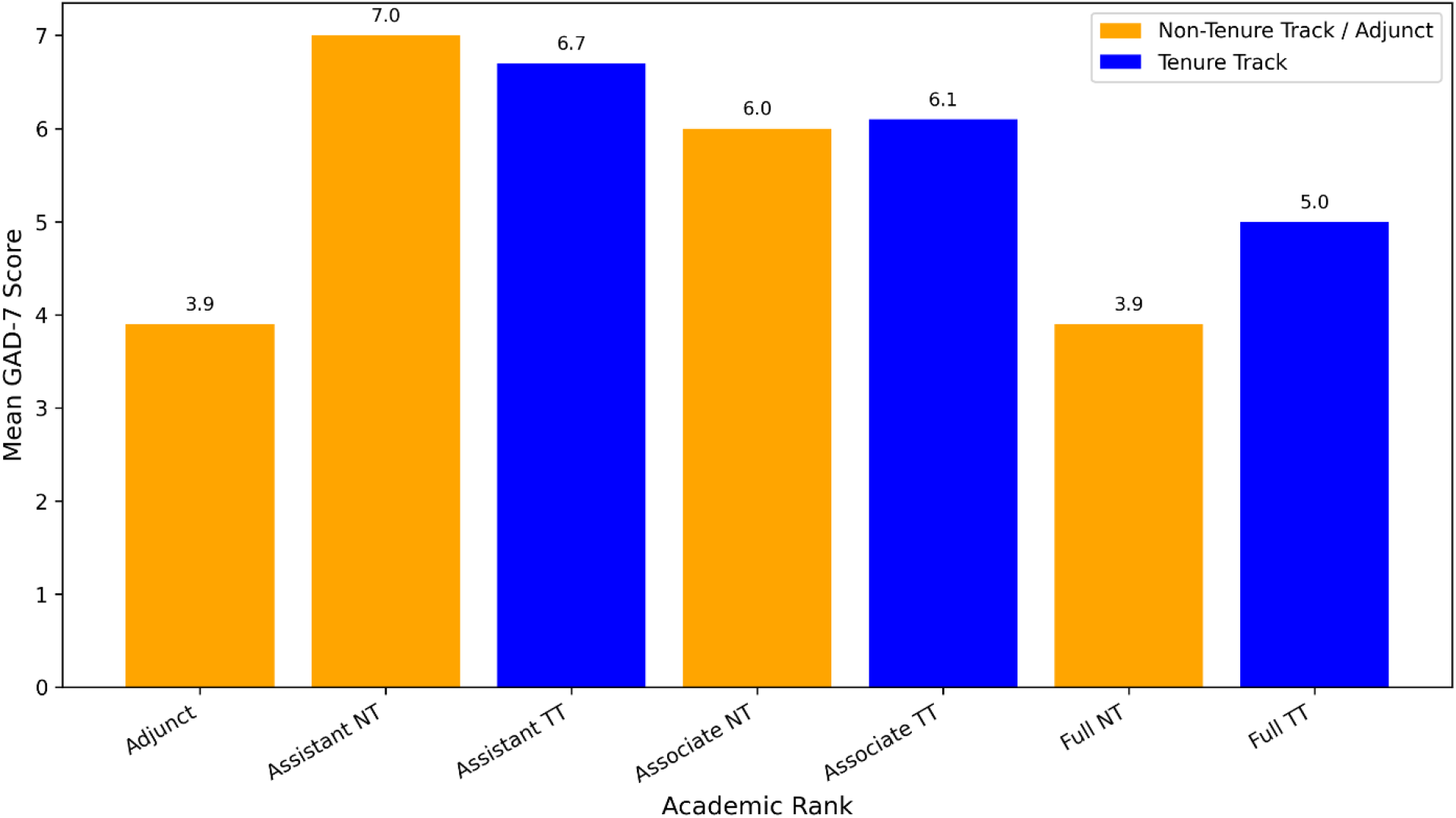
Mean GAD-7 scores by academic rank. Tenure-track (TT) ranks are shown in blue, while non-tenure-track (NT) and adjunct ranks are shown in orange. Assistant Professors (NT and TT) and Associate Professors (NT and TT) report higher mean GAD-7 scores (indicated at the top of each bar) compared to Full Professors and Adjunct or part-time faculty.

### Demographic Predictors

Gender identity was significantly associated with anxiety severity (Figure 2). Gender minorities (self-identified as non-binary or third gender, or those declining to self-identify) reported substantially higher GAD-7 scores compared to female participants, while male participants showed marginally lower GAD-7 scores. These findings suggest that gender identity may be associated with differences in reported anxiety levels, with gender minorities experiencing elevated GAD-7 scores.

**Figure 2.**
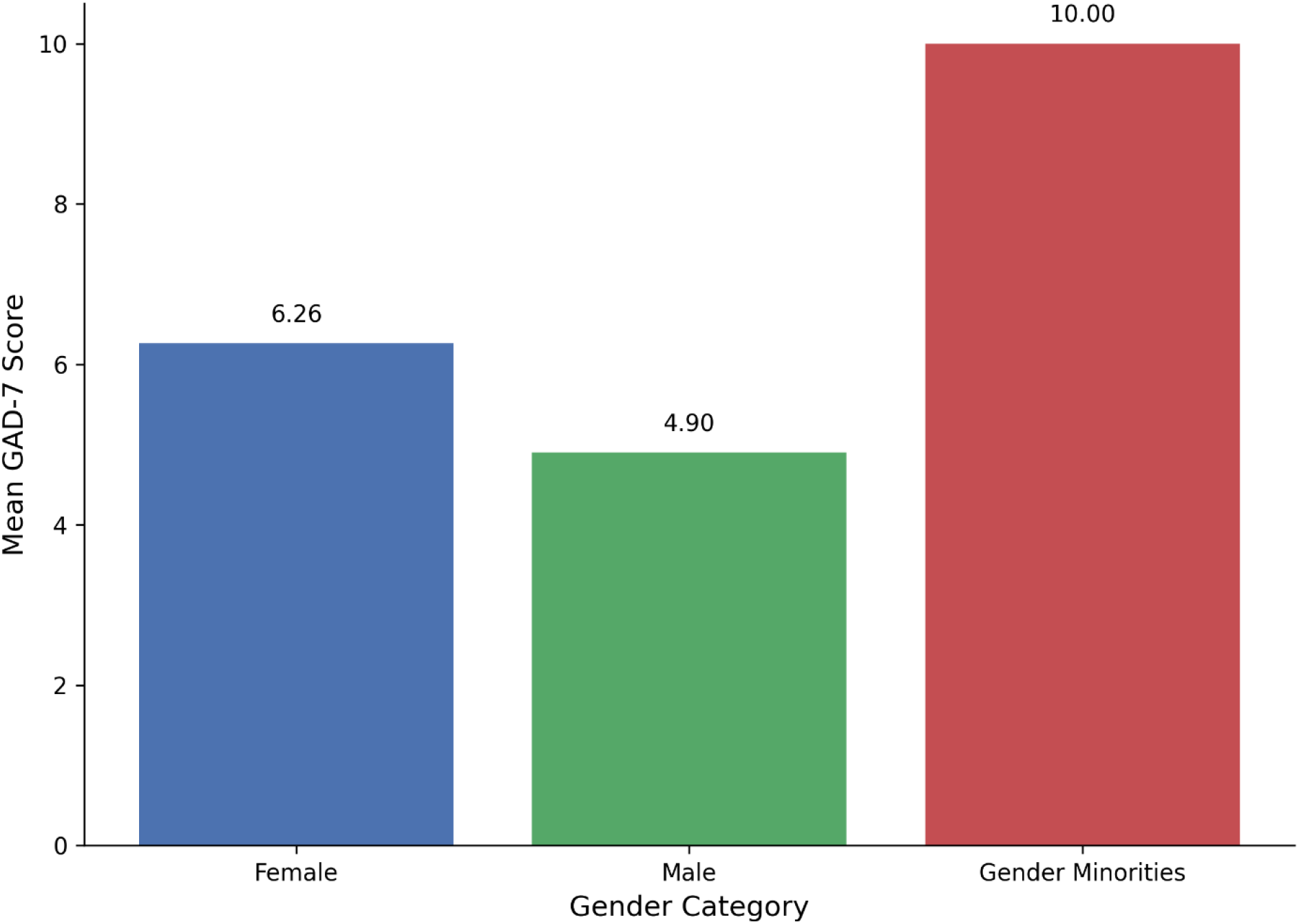
Mean GAD-7 scores by gender category. Gender Minorities include respondents who identified as non-binary/third gender or preferred not to disclose their gender.

A minority of health professions academics have an academic parent, and the vast majority (82.3%) are first-generation academics (Figure 3). Nonetheless, strong majority (89.4%) reported having close family relationships (Figure 3). This illustrates that while close family relationships are common, academic lineage is relatively rare, and most faculty members do not come from academic families. These findings may have implications for understanding access and pathways into academia.

**Figure 3.**
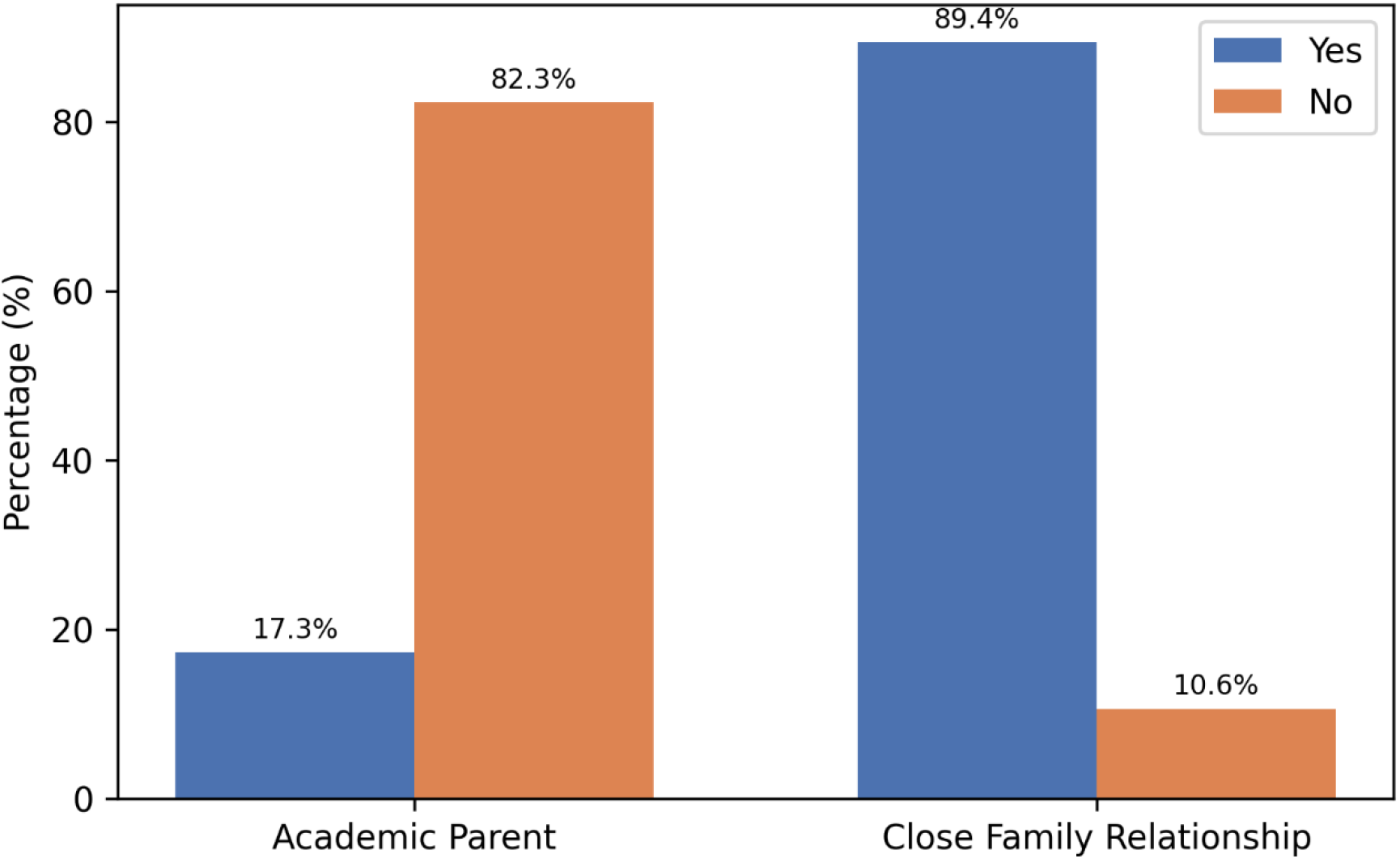
Distribution of academic parent status and close family relationship. Percentage of respondents reporting an academic parent or a close family relationship.

### Effects of Academic Rank and Demographics on Anxiety Severity

A negative binomial regression analysis was conducted to examine predictors of generalized anxiety disorder (GAD) symptom severity among academic professionals (N = 549). The final model included academic rank, gender identity, age, family relationship quality, and academic family background, with statistical significance set at *p* < 0.05. The model was statistically significant overall (LR χ^2^(15) = 65.50, *p* < 0.0001), with a pseudo R^2^ of 0.0209. The dispersion parameter alpha was 0.664 (*p* < 0.001), confirming the appropriateness of the negative binomial model due to overdispersion. All academic ranks showed significantly higher GAD-7 scores compared to adjuncts or part-time faculty (reference group), with the exception of full professors (Table 2). Family relationship quality emerged as a significant predictor (Figure 3). Not having an academic parent was associated with a 31% increase in the expected GAD-7 score compared to those with an academic parent (IRR = 1.31, p = 0.011), which might suggest academic lineage confers protective effects against anxiety, while not having a close family relationship showed a marginally significant 26% increase in GAD-7 score (IRR = 1.26, p = 0.069). Older faculty members, especially those over 65 had much lower anxiety scores compared to younger faculty. For example, faculty aged 65–74 had an 85% lower rate (IRR = 0.15, p = 0.032), and those 75 or older had a 95% lower rate (IRR = 0.05, p = 0.002) compared to the reference group. Other age groups also showed lower rates, though not all were statistically significant. This association could reflect reduced activity, retirement phase, or other factors.

**Table 1.** Respondent Demographics and Characteristics (N = 549)

**Academic and Professional Characteristics**
| **Academic Rank** | **n** | **%** |
| --- | --- | --- |
| Assistant Professor (Non-tenure track) | 153 | 27.9 |
| Associate Professor (Non-tenure track) | 95 | 17.3 |
| Associate Professor (Tenure track) | 76 | 13.8 |
| Assistant Professor (Tenure track) | 76 | 13.8 |
| Full Professor (Tenure track) | 66 | 12.0 |
| Full Professor (Non-tenure track) | 60 | 10.9 |
| Adjunct Professor or Part-time | 23 | 4.2 |
| **Institution Type** | **n** | **%** |
| Research-intensive academic or medical center | 335 | 61.0 |
| Public 4-year university or college | 155 | 28.2 |
| Private 4-year university or college | 38 | 6.9 |
| HBCU or Hispanic-serving institution | 19 | 3.5 |
| Liberal arts college | 2 | 0.4 |
| **Annual Income** | **n** | **%** |
| \$120,000 or more | 410 | 74.7 |
| \$91,000-\$120,000 | 80 | 14.6 |
| \$61,000-\$90,000 | 20 | 3.6 |
| \$31,000-\$60,000 | 4 | 0.7 |
| Prefer not to answer | 35 | 6.4 |

**Demographics**
| **Gender Identity** | **n** | **%** |
| --- | --- | --- |
| Female | 391 | 71.2 |
| Male | 149 | 27.1 |
| Non-binary/third gender | 3 | 0.5 |
| Prefer not to say | 6 | 1.1 |
| **Race/Ethnicity** | **n** | **%** |
| White | 358 | 65.2 |
| Asian or Asian American | 41 | 7.5 |
| Black or African American | 30 | 5.5 |
| Hispanic/Latino | 23 | 4.2 |
| Biracial, multiracial, or mixed race | 11 | 2.0 |
| American Indian or Native American | 1 | 0.2 |
| Other | 85 | 15.5 |
| **Political Orientation** | **n** | **%** |
| Strongly liberal | 192 | 35.0 |
| Strongly liberal | 191 | 34.8 |
| Leaning liberal | 149 | 27.1 |
| Moderate or independent | 106 | 19.3 |

| **Political Orientation** | **n** | **%** |
| --- | --- | --- |
| Leaning conservative | 47 | 8.6 |
| Strongly conservative | 26 | 4.7 |
| Other or prefer not to say | 30 | 5.5 |

Family Characteristics
| **Marital Status** | **n** | **%** |
| --- | --- | --- |
| Married | 424 | 77.2 |
| Single or never married | 69 | 12.6 |
| Divorced | 35 | 6.4 |
| Other/Prefer not to answer | 13 | 2.4 |
| Separated | 4 | 0.7 |
| Widowed | 4 | 0.7 |

| **Has Children** | **n** | **%** |
| --- | --- | --- |
| Yes | 390 | 71.0 |
| No | 149 | 27.1 |
| Missing | 10 | 1.8 |

| **Has Academic parent** | **n** | **%** |
| --- | --- | --- |
| Yes | 95 | 17.3 |
| No | 452 | 82.3 |
| Missing | 2 | 0.4 |

| **Close family relationship** | **n** | **%** |
| --- | --- | --- |
| Yes | 491 | 89.4 |
| No | 58 | 10.6 |

Mental Health Outcomes
| **GAD-7 Severity Level** | **n** | **%** |
| --- | --- | --- |
| Minimal (0-4) | 181 | 38.5 |
| Mild (5-9) | 159 | 33.8 |
| Moderate (10-14) | 92 | 19.6 |
| Severe (15-21) | 38 | 8.1 |

**GAD-7 Total Score:** Mean = 6.97, SD = 5.00 (n = 470)

**Table 2.**
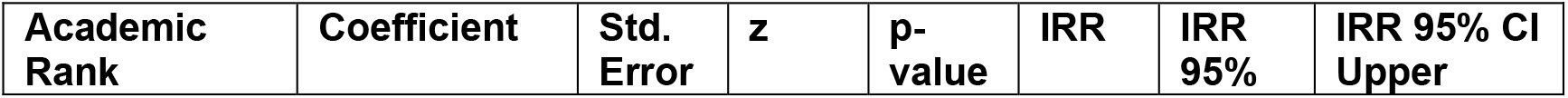
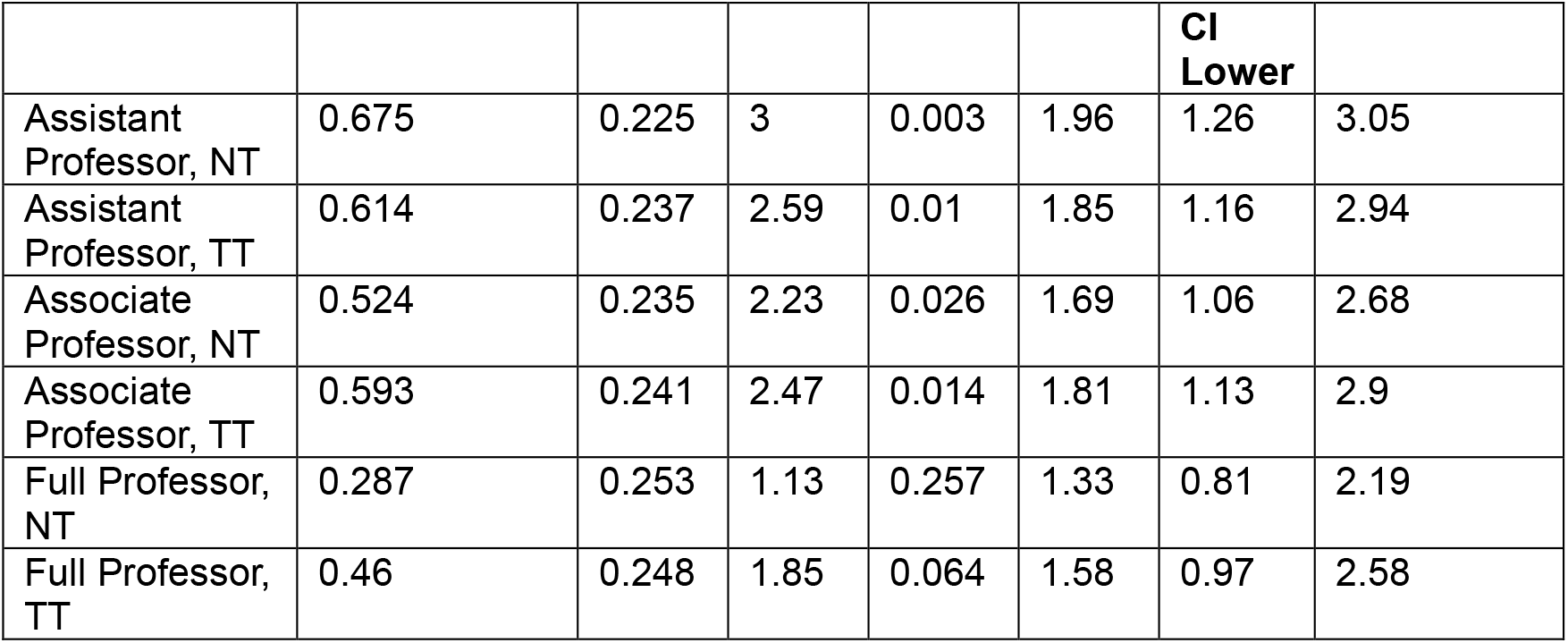
Academic Rank Effects.

## Discussion

This study provides the most comprehensive examination to date of anxiety predictors among academic health professionals, revealing important patterns that challenge some conventional assumptions about academic career stress. Our findings demonstrate that anxiety severity is not uniformly distributed across academic ranks, demographic groups, or social contexts, suggesting the need for targeted interventions. This aligns closely with our earlier work, which examined patterns of anxiety expression among academic faculty and demonstrated that strong familial and parental relationships can significantly mitigate anxiety^8^.

### Academic Rank and Career Stage Effects

Our results reveal a striking pattern whereby assistant professors, particularly those in non-tenure track positions, experience significantly higher anxiety than adjunct or part-time faculty. This finding contradicts common assumptions that job insecurity among adjuncts would translate to higher anxiety levels. Instead, our data suggest that the intense pressures associated with establishing an independent academic career, including research productivity expectations, obtaining extramural funding, and tenure timelines, create particularly high stress environments for junior faculty. The lack of significant anxiety differences among full professors compared to adjunct faculty suggests that anxiety may abate with career advancement and job security, as one may expect. This pattern indicates that interventions should particularly focus on supporting early-career faculty through the most vulnerable phase of their academic careers.

### Age as a Protective Factor

One of the most robust findings in our study was the strong protective effect of age against anxiety symptoms. All age groups showed a progressive and substantially lower anxiety scores than the younger cohort, with the protective effect increasing with age. This pattern may reflect several mechanisms similar to those that reduce anxiety in full professors, such as accumulated coping resources, reduced financial pressures, and differences in career expectations and pressures. The magnitude of this age effect suggests that institutions should pay particular attention to supporting their youngest faculty members, who may lack both life experience and institutional knowledge to navigate academic stressors effectively.

### Demographic Vulnerabilities and Intersectionality

Our findings highlight significant disparities in anxiety levels across demographic groups. Non-binary and third gender respondents reported higher anxiety levels than female participants, with male respondents reporting lowest anxiety scores. This finding underscores the additional stressors faced by women gender minorities in traditional academic environments and highlights the need for inclusive policies and support systems.

### Social and Family Support Systems

Our results confirm the importance of social support systems in buffering against anxiety^8^. Participants with close family relationships and those with academic parents both reported significantly lower anxiety levels. The protective effect of having academic parents may reflect familial factors and environmental advantages, including better understanding of academic culture, expanded professional networks, and potentially reduced financial pressures.

### Implications for Institutional Support

Our findings have several important implications for institutional mental health support strategies. Institutions should develop career-specific targeted support programs for faculty, particularly those in non-tenure track positions. These might include enhanced mentorship programs, clearer advancement pathways, reduced teaching loads during the early career phase, and protected research time. Mental health resources should be designed with awareness of the specific vulnerabilities of younger faculty and women and gender minorities. This might include culturally responsive counseling services, affinity groups, and bias training for leadership. To replicate the effects of close family connections and having an academic lineage, institutions could facilitate family engagement programs, spouse/partner support networks, and community-building activities that extend beyond the immediate academic environment. Rather than focusing solely on individual resilience-building (such as mindfulness or self-care programs), institutions should address systemic factors that contribute to anxiety, including workload management, promotion criteria clarity, and inclusive climate initiatives.

### Limitations

While this study provides valuable insights into anxiety patterns among academic health professionals, several limitations should be noted. The rate of participation in the study may have been affected by response bias since individuals and groups may self-select to participate based on their experience with anxiety and level of satisfaction with the mental health and institutional support available to them. The cross-sectional design prevents causal inferences, and longitudinal studies would better illuminate how anxiety changes throughout academic careers. Additionally, some demographic subgroups had small sample sizes, limiting the generalizability of findings for these populations.

### Future Directions

Future research should investigate the mechanisms underlying the observed demographic and career stage differences in anxiety. Qualitative studies could provide deeper insights into the lived experiences of high-risk groups, while intervention studies could test the effectiveness of targeted support strategies.

### Summary

This study demonstrates that anxiety among academic health professionals follows predictable patterns related to career stage, age, demographic identity, and social support systems. The findings reveal that anxiety severity in academic populations is significantly influenced by career stage, age, gender identity, and family factors. The finding that junior faculty report the highest anxiety levels suggests that the intense pressures of early career development in academic medicine create particularly challenging environments. In addition, younger individuals, gender minorities, and those lacking family support or academic family backgrounds demonstrated elevated anxiety symptoms. These results suggest that targeted interventions addressing the unique stressors faced by vulnerable groups within academic settings may be warranted. Institutions have an opportunity to develop evidence-based, targeted interventions that address both individual vulnerabilities and systemic factors contributing to anxiety among their faculty.

## Data Availability

Due to the IRB
stipulations, the collected data used for this study cannot be shared or made publicly accessible,
however, are available from the corresponding author on reasonable request.

